# Accelerated aging in the brain, epigenetic aging in blood, and polygenic risk for schizophrenia

**DOI:** 10.1101/2020.08.31.20185066

**Authors:** Jalmar Teeuw, Anil Ori, Rachel M. Brouwer, Sonja M.C. de Zwarte, Hugo G. Schnack, Hilleke E. Hulshoff Pol, Roel A. Ophoff

## Abstract

Schizophrenia patients show signs of accelerated aging in cognitive and physiological domains. Both schizophrenia and accelerated aging, as measured by MRI brain images and epigenetic clocks, are correlated with increased mortality. However, the association between these aging measures have not yet been studied in schizophrenia patients.

In schizophrenia patients and healthy subjects, accelerated aging was assessed in brain tissue using a longitudinal MRI (N=715 scans; mean scan interval 3.4 year) and in blood using two epigenetic age clocks (N=172). Differences (‘gaps’) between estimated ages and chronological ages were calculated, as well as the acceleration rate of brain aging. The correlations between these aging measures as well as with polygenic risk scores for schizophrenia (PRS; N=394) were investigated.

Brain aging and epigenetic aging were not significantly correlated. Polygenic risk for schizophrenia was significantly correlated with brain age gap, brain age acceleration rate, and negatively correlated with DNAmAge gap, but not with PhenoAge gap. However, after controlling for disease status and multiple comparisons correction, these effects were no longer significant. Our results imply that the (accelerated) aging observed in the brain and blood reflect distinct biological processes. Our findings will require replication in a larger cohort.

## Introduction

Schizophrenia is a debilitating psychiatric disorder where the patients’ expected lifespan is decreased on average by 15 to 20 years compared to the general population (Laursen et al., 2014; Hjorthøj et al., 2017). This shortened lifespan may be explained in terms of accelerated aging of the body (Kirkpatrick et al., 2008). In two separate studies we have previously reported on accelerated aging of the brain (Schnack et al., 2016) and epigenetic aging in blood (Ori et al., 2019) for schizophrenia patients. However, whether or not these different biomarkers of aging act in concert has noy yet been investigated.

Despite progress in our understanding of neuropsychiatric disorders, the etiology of schizophrenia remains largely unknown. There are several indications of aberrant brain development as early as the fetal period (Debnath et al., 2015; Kim et al., 2015; Faa et al., 2016), with progressive changes of the brain even after the onset of psychosis (van Haren et al., 2008; Hulshoff Pol and Kahn., 2008), which is characteristic for a progressive aging disorder (Olabi et al., 2011). Accelerated biological aging occurs when the rate of biological aging is increased as compared to chronological aging, and may in part explain the increase in mortality rate already at young adult ages within the patient population (Kirkpatrick et al., 2008; Shivakumar et al., 2014; Nguyen et al., 2017; Laursen, et al., 2014). Quantitative assessment of biological aging can be performed using advanced statistical techniques such as machine learning algorithms. These algorithms are trained to discover aging-related patterns in the properties of tissue from a subject or donor (Bzdok, 2017; Cole and Franke, 2017; Jylhävä et al., 2017). Using neuroimaging, the biological age of the brain can be predicted from gray matter distributions (Cole and Franke, 2017; Cole et al., 2017; Valizadeh et al., 2017), white matter properties (Mwangi et al, 2013), or brain-activity related properties (Dosenbach et al, 2010). For patients with schizophrenia, accelerated aging of the brain occurs around the onset of psychosis (Koutsouleris et al., 2014; Schnack et al., 2016; Nenadić et al., 2017; Kaufmann et al., 2019; Jonsson et al., 2019; Kolenic et al., 2018; Chung et al., 2018; Hajek et al., 2019; Shahab et al., 2019) before stabilizing several years after onset (Schnack et al., 2016). Accelerated brain age predicts all-cause mortality (Cole et al., 2018), is highly heritable and has a genetic overlap with common brain disorders, including schizophrenia (Cole et al., 2017; Kaufmann et al., 2019). For biological tissue samples, several molecular and phenotypic biomarkers of aging have been reported for research into proteomic, transcriptomics, metabolomics, telomere length, and DNA methylation levels (Jylhävä et al., 2017; Nguyen et al., 2017). The age of a tissue donor can be reliably estimated based on epigenetic methylation of DNA (Horvath et al., 2012; Horvath, 2013; Hannum et al., 2013; Levine et al., 2018). While no significant accelerated epigenetic aging has previously been observed schizophrenia, neither in post-mortem brain nor blood tissue (Viana et al., 2017; Voisey et al., 2017; McKinney et al., 2017; McKinney et al., 2018), a recent large-scale DNA methylation study now robustly demonstrated that epigenetic age is accelerated in whole blood samples and is strongly correlated with mortality risk (Ori et al., 2019; Higgins-Chen et al., 2020). The study furthermore reports that a subset of cases who carry high schizophrenia polygenic risk present the fastest age acceleration. This raises two intriguing questions; do cases who carry high schizophrenia polygenic risk also present faster brain age acceleration? And is aging in the brain correlated with epigenetic aging in blood? Previously, no significant correlation between brain aging and epigenetic aging was reported in a population study of typical aging elderly subjects (Cole et al., 2018). However, little is known about the interplay between genetics, epigenetics, and brain morphology with regard to accelerated aging in schizophrenia patients. In particular the association between the aging measures based on the brain and blood of schizophrenia patients has not yet been studied.

## Current study

Here we investigated the correlation between brain aging, epigenetic aging, and polygenic risk for schizophrenia within a dataset of schizophrenia patients and healthy control subjects. MRI-derived brain ages were estimated from structural MRI scans using a brain age predictor (Schnack et al., 2016), and epigenetic ages were estimated from whole-blood array-based DNA samples profiles for the DNAmAge clock (Horvath et al., 2012) and the PhenoAge clock (Levine et al., 2018). Genotype-based polygenic risk for schizophrenia was estimated using the schizophrenia GWAS summary statistics of the Psychiatric Genome Consortium (Ripke et al., 2014).

## Materials and methods

### Cohort and sample description

Subjects included in this study were part of two longitudinal schizophrenia cohorts (van Haren et al., 2007; Boos et al., 2012). Brain age in these cohorts has been described before (Schnack et al., 2016) and these cohorts were part of a study on epigenetic aging in schizophrenia (Ori et al., 2019). Here we included unrelated subjects that had imaging data and either epigenetic or genetic data available, resulting in a dataset of 411 unrelated subjects (193 cases, 218 controls, 36% female) of European descent spanning a wide range of the adult lifespan (mean = 32.7 years, range = [16.7 - 67.5] at baseline). For the majority of subjects (57%) longitudinal imaging data was available (up to five scans), with a mean scanning interval of 3.4 years (range [0.9 - 7.0]). All patients met DSM-IV criteria for a nonaffective psychotic disorder (including schizophrenia, schizophreniform disorder or schizoaffective disorder). Written informed consent was obtained from all subjects, and both studies were approved by the Medical Ethics Committee for Research in Humans (METC) of the University Medical Center Utrecht.

**Figure 3.**
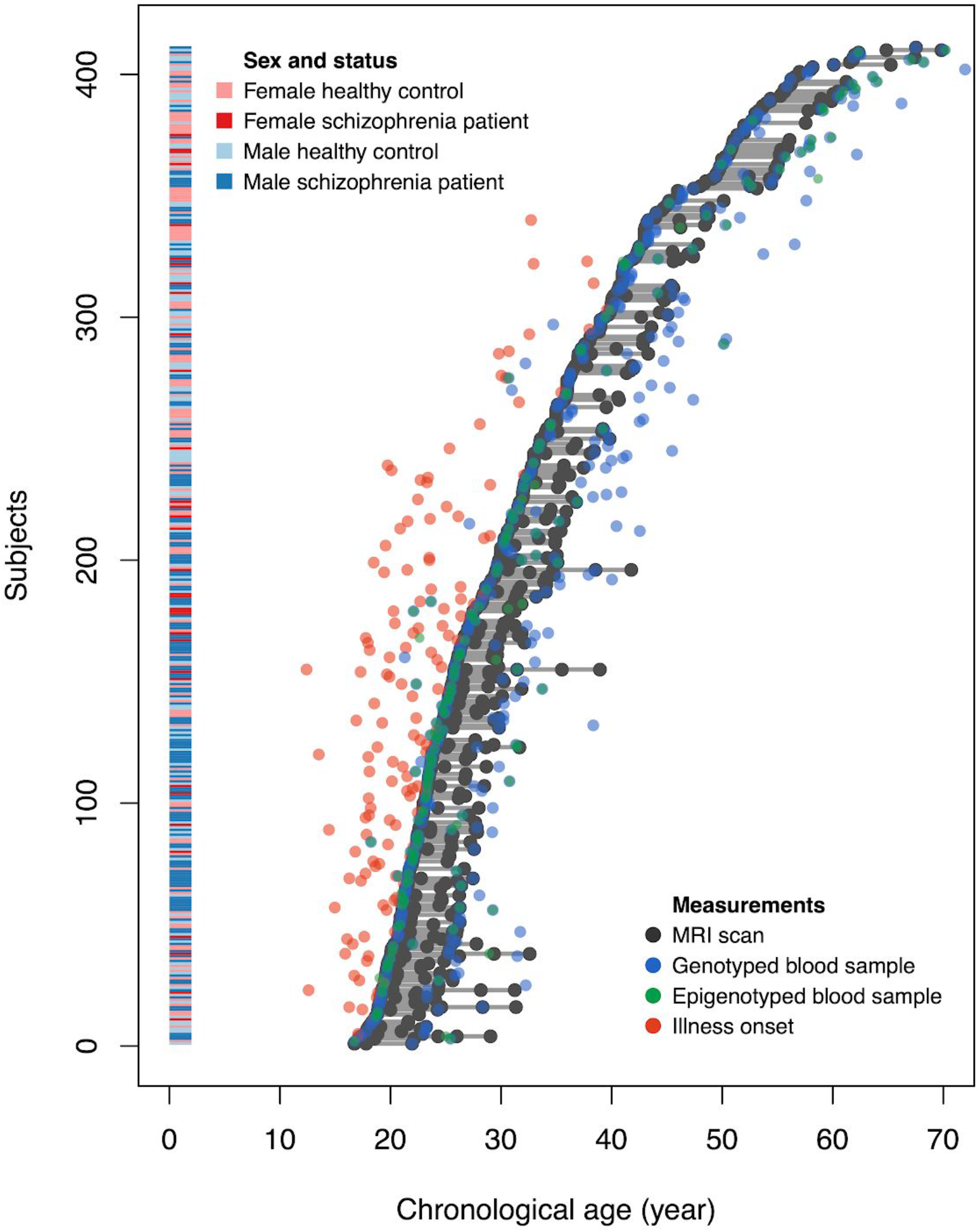
Overview of all subjects with their age at illness onset (not always available for older schizophrenia patients), and age at MRI scans and blood sample acquisition for DNA and epigenetics.

**Table 1.** Demographics table for individuals with data from the three modalities.

| Measure | Population | Subjects with MRI brain age <sup>a</sup> | Subjects with epigenetic ages | Subjects with polygenic risk scores |
| --- | --- | --- | --- | --- |
| Subjects (count) | Total<br>Controls<br>Patients | 411 [715]<br>218 [345]<br>193 [370] | 172<br>63<br>109 | 394<br>212<br>182 |
| Age at baseline (mean±SD) (year) | Total<br>Controls<br>Patients | 32.72±11.41<br>34.92±12.10<br>30.24±10.05 | 32.31±13.01<br>32.62±15.17<br>32.13±11.65 | 35.46±12.47<br>37.55±13.17<br>33.02±11.16 |
| Sex (female:male) | Total<br>Controls<br>Patients | 149:262<br>115:103<br>34:159 | 57:115<br>37:26<br>20:89 | 142:252<br>110:102<br>32:150 |

Abbreviations (in alphabetical order): MRI = magnetic resonance imaging; SD = standard deviation from the mean. <sup>a</sup> Longitudinal imaging data was available for most subjects. Total number of scans between brackets.

### MRI brain age

Structural magnetic resonance imaging (MRI) scans were acquired on a 1.5 tesla Philips scanner with a voxel resolution of 1×1×1.2 mm^3^. Images were processed using a validated in-house image-processing pipeline to produce gray matter density maps in standardized space and used to predict individuals’ brain age. In brief, the predictor uses a model that predicts chronological age based on the weighted sum of whole brain voxel-wise gray matter densities. The model was trained on a sample of healthy control subjects and applied on schizophrenia patients. See (Schnack et al., 2016) for details.

### Blood-based epigenetic aging markers

DNA methylation data was obtained from whole-blood DNA samples using the Illumina Infinium Human Methylation Beadchip technology according to manufacturer’s guidelines. A total of 172 samples were assayed with either the 27K (n=108 samples) or 450K (N=64 samples) platform, which interrogate 27,578 and 485,512 CpG sites across the genome, respectively. These data are a subset of previously published DNAm cohorts (Gene Expression Omnibus (GEO) ID: GSE41037 and GSE41169) for which brain age estimates from MRI scans were available. Blood-based DNAm age was estimated using two different clocks: DNAmAge (Horvath, 2013) and PhenoAge (Levine et al., 2018). These two epigenetic clocks were designed for use with both the 27K and 450K platform allowing us to maximize our sample size. See Supplementary Methods for details.

### Polygenic risk for schizophrenia

Whole-blood DNA samples were processed on Illumina’s HumanOmniExpressExome-8 vl.2 and Illumina’s 550K platform. After quality control (see Supplementary Methods for details), SNPs were imputed on the Michigan server (Das et al., 2016) using the HRC rl.l 2016 reference panel with European samples after phasing with Eagle v2.3. Polygenic risk for schizophrenia was calculated from the SNP data using the schizophrenia GWAS summary statistics of the Psychiatric Genome Consortium excluding Dutch subjects (Ripke et al., 2014). Polygenic scores were calculated using PLINK’s score function at ten GWAS p-value thresholds of significance of the correlation: p < 5×10^−8^, 10^−6^, 10^−4^, 10^−3^, 0.01, 0.05, 0.10, 0.20, 0.5, and 1.0. See Supplementary Methods for details.

Polygenic risk scores were then harmonized to reduce the variation due to acquisition on different platforms through principal component analysis on the full sample. The first principal component contained the majority of the differential disease risk. This component was standardized and used for subsequent analyses, as previously described (Bergen et al., 2019). See Supplementary Methods for details.

### Data preparation

We used a linear mixed-effects regression models to correct the MRI brain age and epigenetic age estimates for regression towards the mean (Le et al., 2018) and differences in acquisition platform within the non-psychiatric controls (**Supplementary Table S1; Supplementary Figure S1**). Age gaps were defined as the difference between the corrected age estimates and the chronological age, and brain age acceleration as the annual rate of change in corrected brain age estimates between consecutive scans. The age gaps and age acceleration were subsequently corrected for possible differences between the sexes by regression of the effect of sex within the healthy control population. The effects of sex were removed from all measures by linear regression regardless of statistical significance and prior to further statistical analyses (**Supplementary Table S2; Supplementary Figure S2**).

### Statistical analysis

First, linear mixed-effects regression models were applied with each of the aging measures as the dependent variable and diagnosis status as fixed effect independent covariate to test for differences between the healthy control and schizophrenia patient groups. The models included random intercepts to account for the repeated measures of the longitudinal MRI scans.

Secondly, the correlations between brain aging (age gap and age acceleration), epigenetic aging (DNAmAge and PhenoAge gap), and polygenic risk for schizophrenia (PRS SCZ) were determined using Spearman’s correlation for all ten pairwise combinations. Since the MRI scans and the blood samples may have been acquired at different visitations, partial correlations were computed between brain age and epigenetic age measures to account for the difference in age of the participant at which the samples were acquired. No correction was applied for correlations involving the polygenic risk for schizophrenia. To assess whether potential correlations were driven by mean differences between patients and controls for both traits, we repeated these analyses correcting for disease status.

For most subjects, more than one MRI scan was available. In the bivariate analyses, the brain age gap from the last MRI scan and the longitudinal brain age acceleration of the first two MRI scans were used. This choice was made based on previous results that show brain age acceleration for schizophrenia patients is maximal around the time of onset (i.e. typically around the acquisition date of the earliest MRI scan) and that its cumulative effect results in a maximal brain age gap for schizophrenia patients 5 years later before stabilizing (Schnack et al., 2016).

A Bonferroni correction was used to account for multiple testing. The corrected significance threshold was set at p=0.05/5=0.01 for group differences and p=0.05/10=0.005 for the tests of pairwise correlations.

## Results

Linear mixed-effects models with random intercepts revealed statistically significant effects of disease status for the MRI-derived brain age gap, longitudinal brain age acceleration, PhenoAge gap, and polygenic risk scores for schizophrenia indicating accelerated age or increased risk for schizophrenia patients, but not for DNAmAge gap (Table 2; Figure 4).

**Table 2.** Group-differences between schizophrenia patients and non**-**patient controls in brain aging, epigenetic aging, and polygenic risk scores for schizophrenia.

| Measure | Healthy controls (Mean±SD) | SCZ Patients (Mean±SD) | Effect of disease status |
| --- | --- | --- | --- |
| MRI brain age gap (years±SD) | -0.06 ± 6.23 | +3.98 ± 7.24 | **+4.03**<br>(**p=2.69e<sup>-9</sup>**)* |
| MRI brain age acceleration (years/year±SD) | 1.01 ± 1.38 | 2.01 ± 2.26 | **+1.00**<br>(**p=7.58e<sup>-5</sup>**)* |
| DNAmAge gap (years±SD) | +0.00 ± 3.56 | -0.52 ± 5.69 | -0.52<br>(p=0.5103) |
| PhenoAge gap (years±SD) | +0.00 ± 6.95 | +2.27 ± 6.74 | **+2.27**<br>(**p=0.0354**) |
| Polygenic risk for schizophrenia (mean ±SD) | +0.00 ± 1.00 | +0.83 ± 0.96 | **+0.83**<br>(**p=6.18e<sup>-16</sup>**)* |

**Figure 4.**
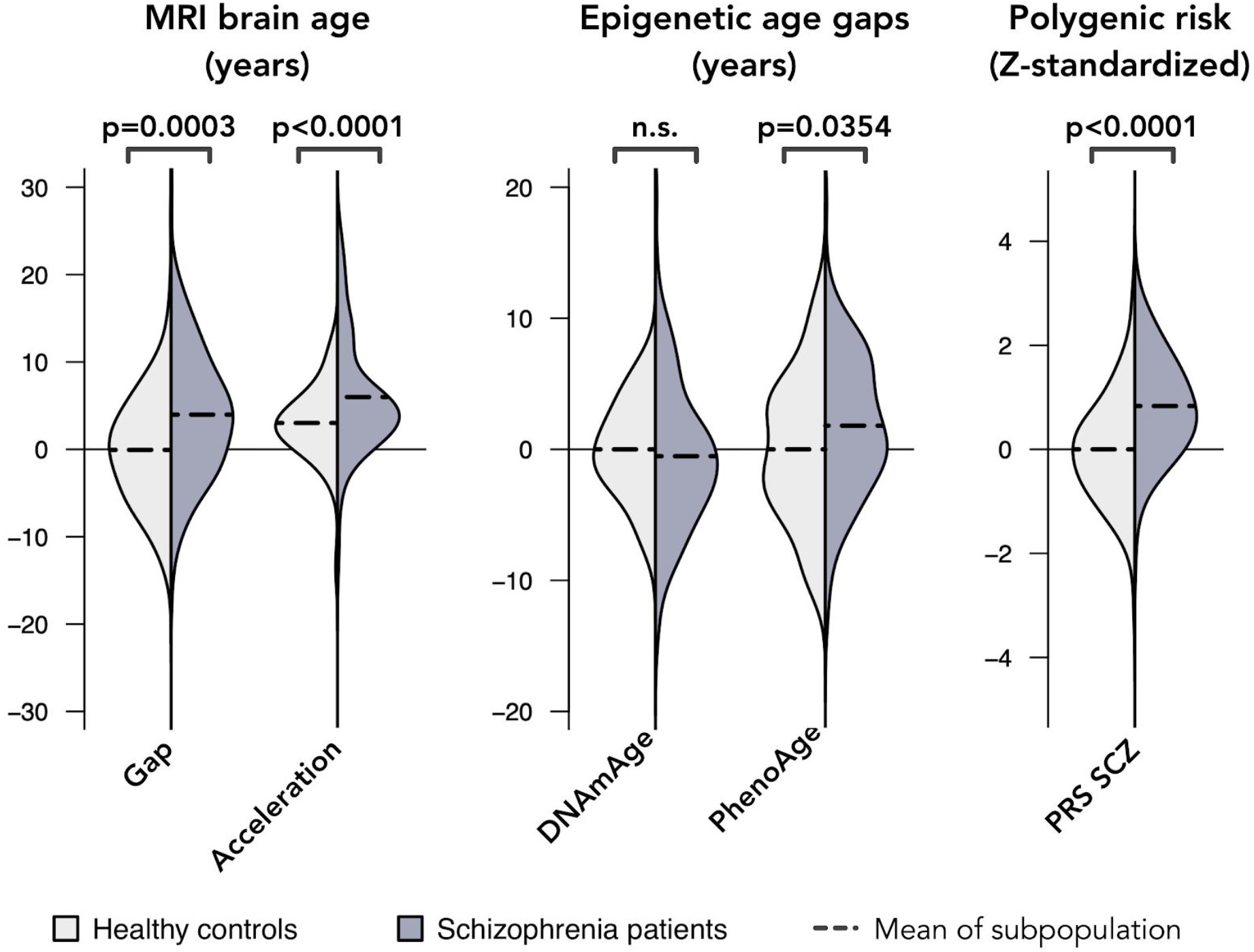
Status effect on the means for MRI-derived brain age gap and acceleration, epigenetic age gaps (DNAmAge and PhenoAge), and polygenic risk for schizophrenia (PRS SCZ). Violin plots show an approximation of the distribution of scores within a subpopulation where density is smoothed by a Gaussian kernel and max height scaled to unit value. Abbreviations (in alphabetical order): PRS = polygenic risk score; SCZ = schizophrenia.

The correlations between measures from the three modalities revealed statistically significant positive correlations between polygenic risk for schizophrenia and MRI-derived brain age gap and brain age acceleration, and a negative correlation between polygenic risk for schizophrenia and DNAmAge gap (**Table 3A; Figure 5**). Within the modalities, there were significant positive correlations between the MRI-derived brain age gap and brain age acceleration, and between the DNAmAge and PhenoAge gaps (**Table 3A; Figure 5**).

After including disease status as a covariate in the partial correlation, the correlation between the MRI-derived brain age gap and acceleration with polygenic risk for schizophrenia were no longer significant (**Table 3B)**.

**Table 3.**
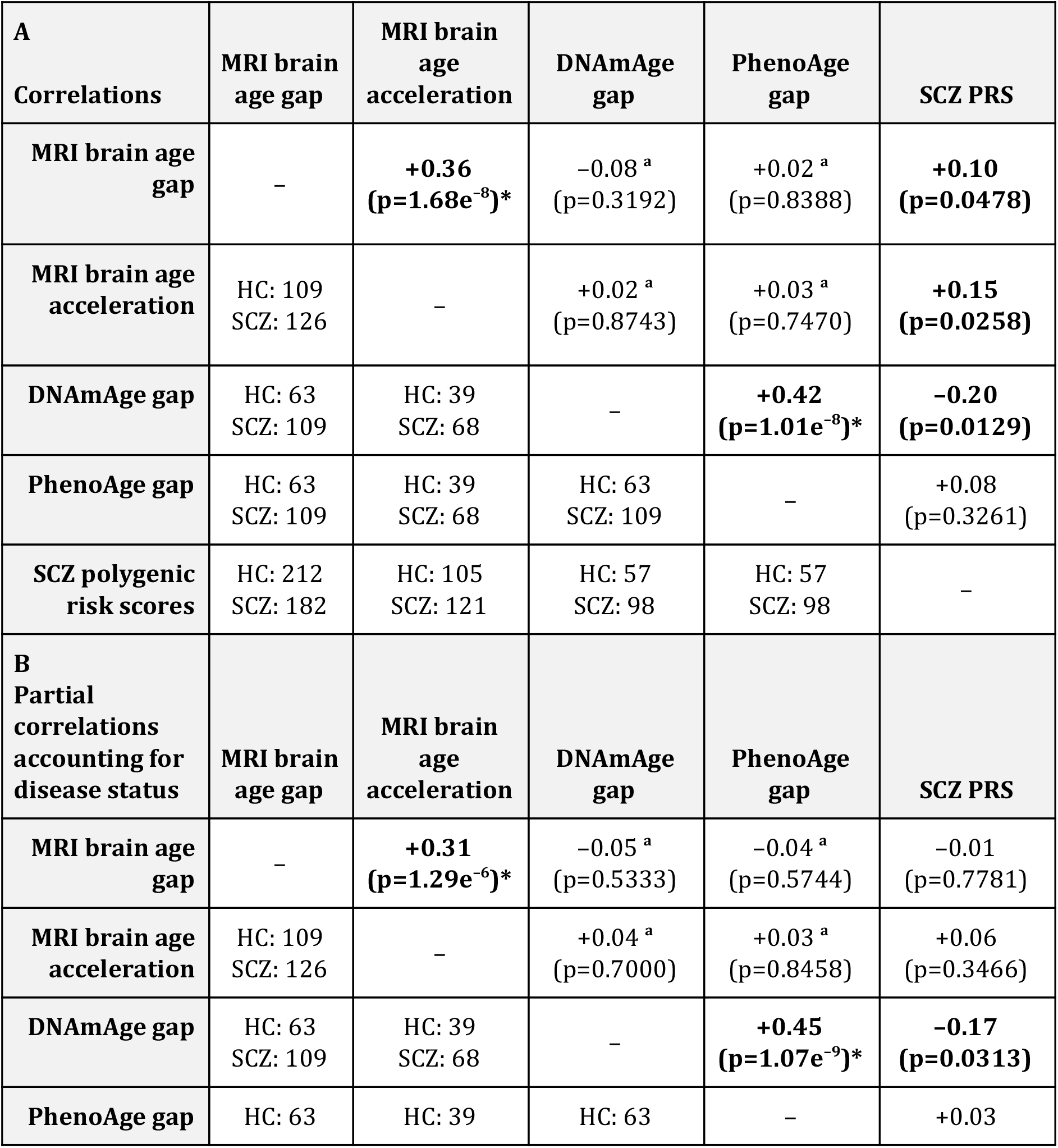
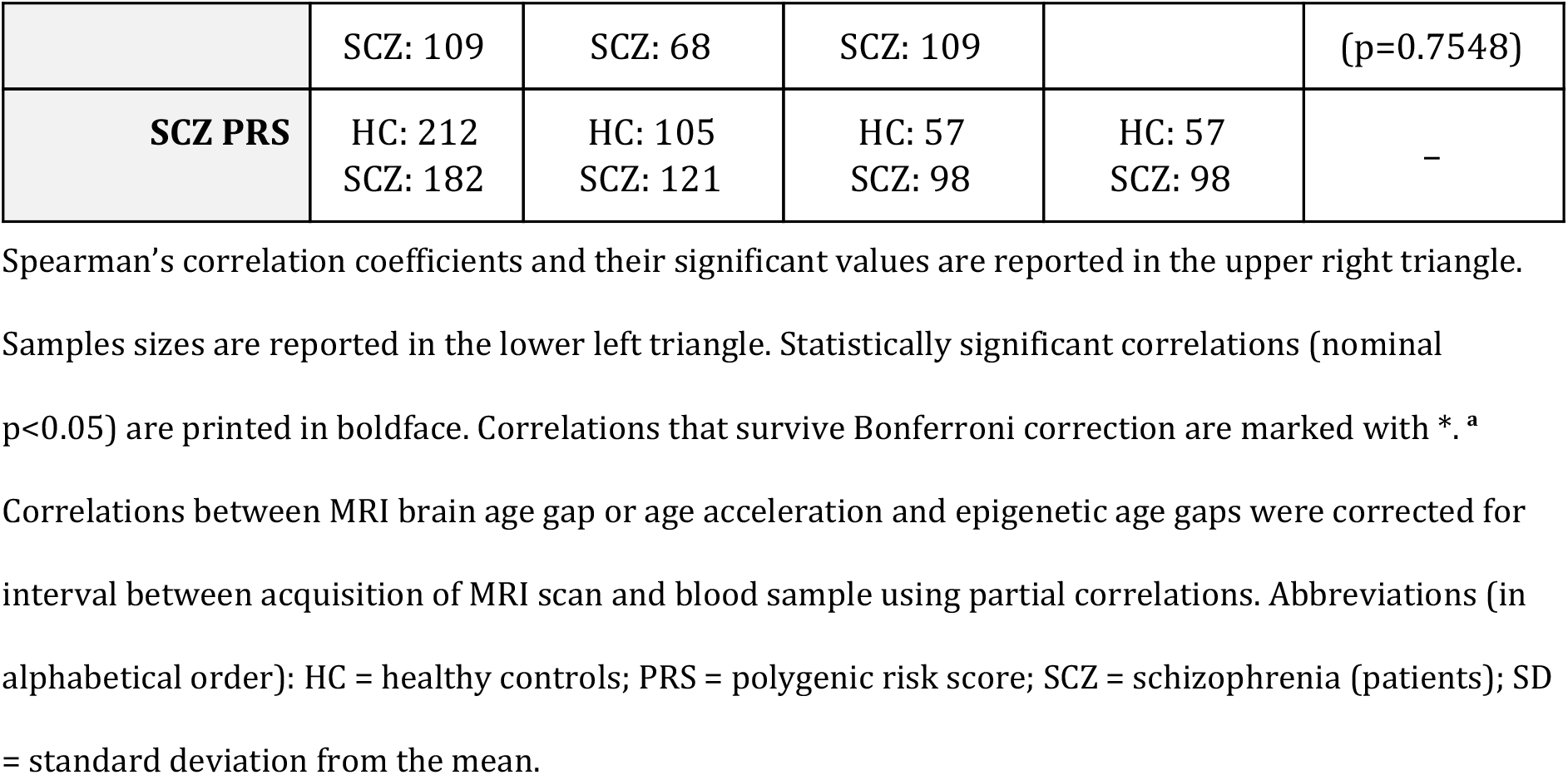
Bivariate analysis between MRI brain age, epigenetic ages, and polygenic risk scores for schizophrenia.

**Figure 5.**
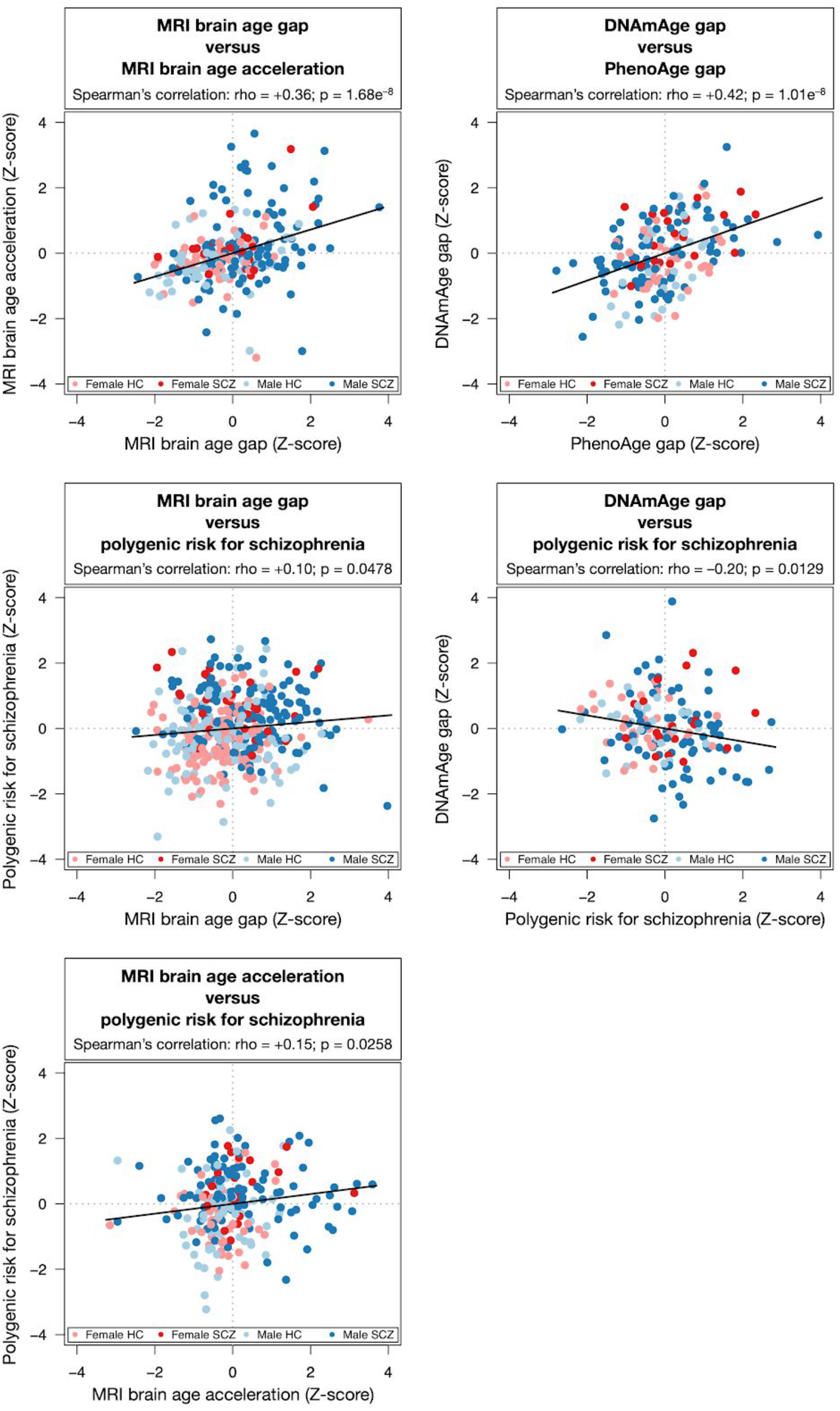
Statistically significant correlations between MRI brain aging, epigenetic aging, and polygenic risk for schizophrenia. Abbreviations (in alphabetical order): HC = healthy controls; SCZ = schizophrenia.

## Discussion

We investigated the correlations between different biological aging markers and with polygenic risk for schizophrenia. We found suggestive evidence of correlations between polygenic risk for schizophrenia with MRI-derived brain aging and with DNAmAge, but not between polygenic risk for schizophrenia and PhenoAge or between brain aging and either epigenetic aging clocks.

### Brain aging in schizophrenia and its correlation with polygenic risk for schizophrenia

Disease status (schizophrenia vs. healthy) had a highly significant effect on brain aging. The age gap, i.e. the difference between estimated age and chronological age, was +4 years in patients, and the brain age acceleration rate was double the rate of healthy controls, consistent with the previously reported results for the broader sample (Schnack et al., 2016) and with other studies reporting accelerated aging of the brain in schizophrenia patients (Koutsouleris et al., 2014; Nenadić et al., 2017; Kaufmann et al., 2019; Jonsson et al., 2019) and in subjects at clinically high-risk for psychosis and first-episode patients (Kolenic et al., 2018; Chung et al., 2018; Hajek et al., 2019; Shahab et al., 2019).

Polygenic risk for schizophrenia (PRS), as expected, was significantly higher in the patients as compared to the control subjects. We found nominal significant correlations between polygenic risk for schizophrenia and MRI-derived brain age gap *(rho* = +0.10) and longitudinal acceleration *(rho* = +0.15). These correlations were largely moderated by disease status, indicating that cases who carry higher polygenic risk for schizophrenia display faster brain age acceleration. Our results are in line with recent work that observed an overlap between common genetic variants associated with brain aging and common variants associated with schizophrenia in the population (Kaufmann et al., 2019). Here, we observe a direct correlation between brain aging and schizophrenia polygenic risk within individuals. While these findings provide evidence for a shared mechanism between genetic risk of schizophrenia and aging in the brain, replication and further work in a larger sample should be a first priority for future work. This study nevertheless reports the first efforts and value of such analyses. Structural brain aging could thus be used as an intermediate phenotype for psychosis (Palaniyappan et al., 2017; Dukart et al., 2017) and may show promise in predicting transition to psychosis in at-risk populations (Koutsouleris et al., 2014).

### Epigenetic aging in schizophrenia and its correlations with polygenic risk for schizophrenia

We found that epigenetic age was significantly accelerated by +2.3 year in schizophrenia patients as compared to healthy controls for the PhenoAge clock, but not for the DNAmAge clock. Previous reports suggest no accelerated epigenetic aging or association with premature mortality for schizophrenia in blood or post-mortem brain samples (McKinney et al., 2017; Voisey et al., 2017; McKinney et al., 2018; Kowalec et al., 2019), with some exceptions (Okazaki et al., 2019; Ori et al., 2019; Higgins-Chen, 2020).

In addition, we found a significant negative correlation *(rho* = −0.20) between polygenic risk for schizophrenia and DNAmAge gap regardless of diagnosis status, but not for PhenoAge gap *(rho* = +0.08; [n.s.]). However, the negative correlation for DNAmAge gap did not survive correction for multiple testing and the correlation was not significant in our previous analysis in a larger sample (Ori et al., 2019). Moreover, the previous analysis in the broader sample did find a significant correlation between polygenic risk for schizophrenia and PhenoAge gap that was age- and sex-specific, with female patients above age 36 showing an increase in PhenoAge gap of +3 years (Ori et al., 2019). However, the current sample is too small to stratify by age group or sex. In another study, polygenic risk for schizophrenia was associated with mortality predictions based on the PhenoAge clock and suicidal behavior (Laursen et al., 2017). These contradicting results warrant caution in interpreting our findings and emphasizes the need for follow-up studies in larger samples.

Accelerated epigenetic aging is heritable (Marioni et al., 2015; Li et al., 2015), with an important role for the TERT locus related to telomerase and aging and nine other loci related to metabolism and immune system pathways (Lu et al., 2017; Gibson et al., 2019). No overlap between genetic variants identified for schizophrenia and epigenetic aging is reported in a relatively small sample (Lu et al., 2018). There is an indication for colocalization of genetic and epigenetic loci implicated in schizophrenia (Hannon et al., 2016), however, epigenetic loci used to predict epigenetic age do not overlap with known epigenetic loci implicated in schizophrenia (Mill et al., 2008; Hannon et al., 2016). Instead, it is possible that the correlation between epigenetic age and polygenic risk for schizophrenia is mediated by other factors, e.g. a shared pathway that increases risk of early mortality (Marioni et al., 2015; Levine et al, 2018; Laursen et al., 2014) such a genetic predisposition to smoking (Boardman et al., 2010) or stressful life events (Wolf et al., 2018).

### Brain aging and its correlation with epigenetic aging

We did not find any significant correlation between the MRI-derived brain age gap or age acceleration with the DNAmAge or PhenoAge gap (range of *rho* = [-0.08; +0.03]; nominal *p >* 0.32). An absence of a correlation between the MRI-derived brain age gap and DNAmAge gap in blood samples has previously been reported in elderly subjects (Cole et al., 2018). Here we complement the previous finding by not only replicating the null result for the DNAmAge clock (that is a reliable predictor of chronological age regardless of tissue type or disease), but also investigating the correlation between MRI brain aging and the PhenoAge clock in blood (which might be more sensitive to aberrant biological aging due to the inclusion of extrinsic factors more representative of apparent phenotypic aging) (Levine et al., 2018). However, the correlations remained absent despite the effects of disease status and the correlations with polygenic risk for schizophrenia in the individual aging measures. As previously reported in broader samples, epigenetic aging in schizophrenia shows an age- and sex-specific effect in a larger study that included this cohort (Ori et al., 2019), and accelerated aging of the brain is already present in the first years following the onset of psychosis before it stabilizes several years afterward (Schnack et al., 2016). The absence of a correlation between these two aging measures might be due to distinct aging processes. Epigenetic aging, in particular the DNAmAge clock, is a measure of cellular aging rather than cellular senescence (Lowe et al., 2016; Kabacik et al., 2018). In contrast, the aging of the brain, in our study reflecting decreases in gray matter tissue, is likely due to cell senescence rather than cellular aging (Fernandez-Egea, 2017), and it reflects changes in the morphology of the cells or composition of the neuropil. This argument is used to explain the absence of accelerated epigenetic aging in post-mortem samples of the brains of schizophrenia patients (McKinney et al., 2017; Voisey et al., 2017; McKinney et al., 2018). The possibility of two independent aging processes was previously suggested given the lack of correlation between the MRI-derived and epigenetic age gaps and the fact that combining information from the two ‘clocks’ improved mortality predictions (Cole et al., 2018). The absence of a correlation with PhenoAge gap, one that takes into account extrinsic factors of typical aging, from our results affirms the conclusion that aging of the brain and epigenetic aging in blood might be two distinct processes in the aetiology of schizophrenia, despite their commonality in predicting mortality (Cole et al., 2018; Marioni et al., 2015; Chen et al., 2016). A similar conclusion on the dissociation between brain aging and epigenetic aging can be concluded for related psychiatric disorders based on the reports from several independent studies. For bipolar disorder, accelerated epigenetic aging (Nenadić et al., 2017) but not aging of the brain (Fries et al., 2017; Shahab et al., 2019; Nenadić et al., 2017) has been reported, although lithium use may have confounded these results, since patients who were not treated with lithium have been found to show increased brain age (Van Gestel et al., 2019). For major depressive disorder, a large (N=1689) international multicenter study (Han et al., 2020) has found accelerated aging of the brain, but results are inconclusive within smaller samples (Koutsouleris et al, 2014; Besteher et al, 2019; Kaufmann et al, 2019). Epigenetic aging in blood (Han et al., 2018) but not epigenetic aging in post-mortem brain samples (Li et al., 2018) has been reported. These studies suggest the possibility for distinct aging processing for brain tissue and blood across psychiatric disorders.

### Limitations and future directions

There are a few limitations to this study that should be taken into account. First, the sample size of this study, whilst large for a longitudinal neuroimaging study, is very modest for a genetic or epigenetic study. Secondly, the cross-sectional design for epigenetics limits our ability to detect a possible age acceleration rate in the blood and its correlation to accelerating brain age (Nelson et al., 2019). Depending on the time lag between illness onset and accelerated epigenetic aging, and because of the fact that most of the blood sample were acquired at baseline, the effects of the disease on DNA methylation may have yet to occur, especially in the younger adolescent population when onset of psychosis typically occurs (Paus et al., 2008). Future studies, measuring both brain aging and epigenetic aging in large longitudinal studies should further elucidate the possible (dynamic) relationships between these different measures of biological aging.

## Data Availability

Requests for access to the data and code used in this analysis should be directed to the corresponding author.

## References

Alnæs D, Kaufmann T, van der Meer D, Córdova-Palomera A, Rokicki J, Moberget T, Bettella F, Agartz I, Barch DM, Bertolino A, Brandt CL, Cervenka S, Djurovic S, Doan NT, Eisenacher S, Fatouros-Bergman H, Flyckt L, Di Giorgio A, Haatveit B, Jönsson EG, KaSP Consortium, Kirsch P, Lund MJ, Meyer-Lindenberg A, Pergola G, Schwarz E, Smeland OB, Quarto T, Zink M, Andreassen OA, Westlye LT. 2019. The dark side of the mean: brain structural heterogeneity in schizophrenia and its polygenic risk. bioRxiv. 8:12.

Arion D, Corradi JP, Tang S, Datta D, Boothe F, He A, Cacace AM, Zaczek R, Albright CF, Tseng G, Lewis DA. 2015. Distinctive transcriptome alterations of prefrontal pyramidal neurons in schizophrenia and schizoaffective disorder. Mol Psychiatry. 20:1397–1405.

Bakulski KM, Halladay A, Hu VW, Mill J, Fallin MD. 2016. Epigenetic Research in Neuropsychiatric Disorders: the “Tissue Issue”. Curr Behav Neurosci Rep. 3:264–274.

Bayer TA, Falkai P, Maier W. 1999. Genetic and non-genetic vulnerability factors in schizophrenia: The basis of the “Two hit hypothesis.” Journal of Psychiatric Research. 33:543–548.

Belsky DW, Moffitt TE, Cohen AA, Corcoran DL, Levine ME, Prinz JA, Schaefer J, Sugden K, Williams B, Poulton R, Caspi A. 2018. Eleven Telomere, Epigenetic Clock, and Biomarker-Composite Quantifications of Biological Aging: Do They Measure the Same Thing? Am J Epidemiol. 187:1220–1230.

Bergen SE, Ploner A, Howrigan D, O’Donovan MC, Smoller JW, Sullivan PF, Sebat J, Neale B, Kendler KS. 2019. Joint contributions of rare copy number variants and common SNPs to risk for schizophrenia. AJP. 176:29–35.

Besteher B, Gaser C, Nenadić I. Machine-learning based brain age estimation in major depression showing no evidence of accelerated aging. Psychiatry Research: Neuroimaging. 2019 Aug 30;290:1–4.

Binder AM, Corvalan C, Mericq V, Pereira A, Santos JL, Horvath S, Shepherd J, Michels KB. 2018. Faster ticking rate of the epigenetic clock is associated with faster pubertal development in girls. Epigenetics. 13:85–94.

Boardman JD, Blalock CL, Pampel FC. 2010. Trends in the genetic influences on smoking. J Health Soc Behav. 51:108–123.

Bocklandt S, Lin W, Sehl ME, Sánchez FJ, Sinsheimer JS, Horvath S, Vilain E. 2011. Epigenetic predictor of age. PLoS ONE. 6:e14821.

Boos HB, Cahn W, van Haren NE, Derks EM, Brouwer RM, Schnack HG, Hulshoff Pol HE, Kahn RS. 2012. Focal and global brain measurements in siblings of patients with schizophrenia. Schizophr Bull 38:814–825.

Bzdok D. 2017. Classical statistics and statistical learning in imaging neuroscience. Front Neurosci. 11:543.

Cariaga-Martinez A, Alelú-Paz R. 2016. False data, positive results in neurobiology: Moving beyond the epigenetics of blood and saliva samples in mental disorders. J Negat Results Biomed. 15:21.

Cevenini E, Invidia L, Lescai F, Salvioli S, Tieri P, Castellani G, Franceschi C. 2008. Human models of aging and longevity. Expert Opin Biol Ther. 8:1393–1405.

Chen BH, Marioni RE, Colicino E, Peters MJ, Ward-Caviness CK, Tsai P-C, Roetker NS, Just AC, Demerath EW, Guan W, Bressler J, Fornage M, Studenski S, Vandiver AR, Moore AZ, Tanaka T, Kiel DP, Liang L, Vokonas P, Schwartz J, Lunetta KL, Murabito JM, Bandinelli S, Hernandez DG, Melzer D, Nalls M, Pilling LC, Price TR, Singleton AB, Gieger C, Holle R, Kretschmer A, Kronenberg F, Kunze S, Linseisen J, Meisinger C, Rathmann W, Waldenberger M, Visscher PM, Shah S, Wray NR, McRae AF, Franco OH, Hofman A, Uitterlinden AG, Absher D, Assimes T, Levine ME, Lu AT, Tsao PS, Hou L, Manson JE, Carty CL, LaCroix AZ, Reiner AP, Spector TD, Feinberg AP, Levy D, Baccarelli A, van Meurs J, Bell JT, Peters A, Deary IJ, Pankow JS, Ferrucci L, Horvath S. 2016. DNA methylation-based measures of biological age: meta-analysis predicting time to death. Aging (Albany NY). 8:1844–1865.

Chinta SJ, Woods G, Rane A, Demaria M, Campisi J, Andersen JK. 2015. Cellular senescence and the aging brain. Exp Gerontol. 68:3–7.

Chung Y, Addington J, Bearden CE, Cadenhead K, Cornblatt B, Mathalon DH, McGlashan T, Perkins D, Seidman LJ, Tsuang M, Walker E, Woods SW, McEwen S, van Erp TGM, Cannon TD. 2018. Use of machine learning to determine deviance in neuroanatomical maturity associated with future psychosis in youths at clinically high risk. JAMA Psychiatry. 75:960–968.

Cole JH, Franke K. 2017. Predicting Age Using Neuroimaging: Innovative Brain Ageing Biomarkers. Trends Neurosci. 40:681–690.

Cole JH, Marioni RE, Harris SE, Deary IJ. 2019. Brain age and other bodily “ages”: implications for neuropsychiatry. Mol Psychiatry. 24:266–281.

Cole JH, Poudel RPK, Tsagkrasoulis D, Caan MWA, Steves C, Spector TD, Montana G. 2017. Predicting brain age with deep learning from raw imaging data results in a reliable and heritable biomarker. Neuroimage. 163:115–124.

Cole JH, Ritchie SJ, Bastin ME, Valdés Hernández MC, Muñoz Maniega S, Royle N, Corley J, Pattie A, Harris SE, Zhang Q, Wray NR, Redmond P, Marioni RE, Starr JM, Cox SR, Wardlaw JM, Sharp DJ, Deary IJ. 2018. Brain age predicts mortality. Mol Psychiatry. 23:1385–1392.

Das S, Forer L, Schonherr S, Sidore C, Locke AE, Kwong A, Vrieze SI, Chew EY, Levy S, McGue M, Schlessinger D, Stambolian D, Loh P-R, Iacono WG, Swaroop A, Scott LJ, Cucca F, Kronenberg F, Boehnke M, Abecasis GR, Fuchsberger C. 2016. Next-generation genotype imputation service and methods. Nat Genet. 48:1284–1287.

de Nooij L, Harris MA, Hawkins EL, Shen X, Clarke T-K, Chan SWY, Ziermans TB, McIntosh AM, Whalley HC. 2019. Longitudinal trajectories of brain age in young individuals at familial risk of mood disorder. bioRxiv. 27:3.

Debnath M, Venkatasubramanian G, Berk M. 2015. Fetal programming of schizophrenia: Select mechanisms. Neuroscience and Biobehavioral Reviews. 49:90–104.

Dosenbach NU, Nardos B, Cohen AL, et al: Prediction of individual brain maturity using fMRI. Science 2010; 329:1358–1361.

Dukart J, Smieskova R, Harrisberger F, Lenz C, Schmidt A, Walter A, Huber C, Riecher-Rossler A, Simon A, Lang UE, Fusar-Poli P, Borgwardt S. 2017. Age-related brain structural alterations as an intermediate phenotype of psychosis. JPN. 42:307–319.

Faa G, Manchia M, Pintus R, Gerosa C, Marcialis MA, Fanos V. 2016. Fetal programming of neuropsychiatric disorders. Birth Defects Res C Embryo Today. 108:207–223.

Farré P, Jones MJ, Meaney MJ, Emberly E, Turecki G, Kobor MS. 2015. Concordant and discordant DNA methylation signatures of aging in human blood and brain. Epigenetics Chromatin. 8:19.

Fernandez-Egea E, Kirkpatrick B. 2017. Correspondence regarding two recent publications in npj:schizophrenia about DNAm and accelerated aging in schizophrenia, npj Schizophrenia. 3:38.

Fries GR, Bauer IE, Scaini G, Wu M-J, Kazimi IF, Valvassori SS, Zunta-Soares G, Walss-Bass C, Soares JC, Quevedo J. 2017. Accelerated epigenetic aging and mitochondrial DNA copy number in bipolar disorder. Translational Psychiatry. 7:1283.

Gao X, Zhang Y, Breitling LP, Brenner H. 2016. Tobacco smoking and methylation of genes related to lung cancer development. Oncotarget. 7:59017–59028.

Glausier JR, Lewis DA. 2013. Dendritic spine pathology in schizophrenia. Neuroscience. 251:90–107.

Hajek T, Franke K, Kolenic M, Capkova J, Matejka M, Propper L, Uher R, Stopkova P, Novak T, Paus T, Kopecek M, Spaniel F, Alda M. 2019. Brain Age in Early Stages of Bipolar Disorders or Schizophrenia. Schizophr Bull. 45:190–198.

Han LKM, Aghajani M, Clark SL, Chan RF, Hattab MW, Shabalin AA, Zhao M, Kumar G, Xie LY, Jansen R, Milaneschi Y, Dean B, Aberg KA, van den Oord EJCG, Penninx BWJH. 2018. Epigenetic aging in major depressive disorder. AJP. 175:774–782.

Han LKM, Dinga R, Hahn T, et al. 2020. Brain aging in major depressive disorder: results from the ENIGMA major depressive disorder working group. Mol Psychiatry. 2020 May 18.

Hannon E, Dempster E, Viana J, Burrage J, Smith AR, Macdonald R, St Clair D, Mustard C, Breen G, Therman S, Kaprio J, Toulopoulou T, Hulshoff Pol HE, Bohlken MM, Kahn RS, Nenadić I, Hultman CM, Murray RM, Collier DA, Bass N, Gurling H, McQuillin A, Schalkwyk L, Mill J. 2016. An integrated genetic-epigenetic analysis of schizophrenia: Evidence for co-localization of genetic associations and differential DNA methylation. Genome Biol. 17:176.

Hannum G, Guinney J, Zhao L, Zhang L, Hughes G, Sadda SV, Klotzle B, Bibikova M, Fan J-B, Gao Y, Deconde R, Chen M, Rajapakse I, Friend S, Ideker T, Zhang K. 2013. Genome-wide Methylation Profiles Reveal Quantitative Views of Human Aging Rates. Mol Cell. 49:359–367.

He J, Kong J, Tan Q-R, Li X-M. 2009. Neuroprotective effect of atypical antipsychotics in cognitive and non-cognitive behavioral impairment in animal models. Cell Adh Migr. 3:129–137.

Higgins-Chen, A., Boks, M., Vinkers, C., Kahn, R., Levine, M. (2020). Schizophrenia and Epigenetic Aging Biomarkers: Increased Mortality, Reduced Cancer Risk, and Unique Clozapine Effects Biological Psychiatry 88(3), 224-235. https://dx.doi.org/10.1016/j.biopsych.2020.01.025

Hillary RF, Stevenson AJ, Cox SR, McCartney DL, Harris SE, Seeboth A, Higham J, Sproul D, Taylor AM, Redmond P, Corley J, Pattie A, Valdés Hernández MDC, Muñoz-Maniega S, Bastin ME, Wardlaw JM, Horvath S, Ritchie CW, Spires-Jones TL, McIntosh AM, Evans KL, Deary IJ, Marioni RE. 2019. An epigenetic predictor of death captures multi-modal measures of brain health. bioRxiv. 10:573.

Horvath S. 2013. DNA methylation age of human tissues and cell types. Genome Biol. 14:R115-R120.

Horvath S, Gurven M, Levine ME, Trumble BC, Kaplan H, Allayee H, Ritz BR, Chen B, Lu AT, Rickabaugh TM, Jamieson BD, Sun D, Li S, Chen W, Quintana-Murci L, Fagny M, Kobor MS, Tsao PS, Reiner AP, Edlefsen KL, Absher D, Assimes TL. 2016. An epigenetic clock analysis of race/ethnicity, sex, and coronary heart disease. Genome Biol. 17:171.

Horvath S, Zhang Y, Langfelder P, Kahn RS, Boks MPM, van Eijk K, van den Berg LH, Ophoff RA. 2012. Aging effects on DNA methylation modules in human brain and blood tissue. Genome Biol. 13:R97-18.

Hulshoff Pol HE, Kahn RS. 2008. What happens after the first episode? A review of progressive brain changes in chronically ill patients with schizophrenia. Schizophr Bull. 34:354–366.

Jaffe AE, Gao Y, Deep-Soboslay A, Tao R, Hyde TM, Weinberger DR, Kleinman JE. 2015. Mapping DNA methylation across development, genotype and schizophrenia in the human frontal cortex. Nat Neurosci. 19:40–47.

Jeste DV, Wolkowitz OM, Palmer BW. 2011. Divergent trajectories of physical, cognitive, and psychosocial aging in schizophrenia. Schizophr Bull. 37:451–455.

Jonsson BA, Bjornsdottir G, Thorgeirsson TE, Ellingsen LM, Bragi Walters G, Gudbjartsson DF, Stefansson H, Stefansson K, Ulfarsson MO. 2019. Deep learning based brain age prediction uncovers associated sequence variants. bioRxiv. 475:S2.

Jylhävä J, Pedersen NL, Hägg S. 2017. Biological Age Predictors. EBioMedicine. 21:29–36.

Kabacik S, Horvath S, Cohen H, Raj K. 2018. Epigenetic ageing is distinct from senescence-mediated ageing and is not prevented by telomerase expression. Aging (Albany NY). 10:2800–2815.

Kaufmann T, van der Meer D, Doan NT, Schwarz E, Lund MJ, Agartz I, Alnæs D, Barch DM, Baur-Streubel R, Bertolino A, Bettella F, Beyer MK, Bøen E, Borgwardt S, Brandt CL, Buitelaar J, Celius EG, Cervenka S, Conzelmann A, Córdova-Palomera A, Dale AM, de Quervain DJF, Carlo PD, Djurovic S, Dørum ES, Eisenacher S, Elvsåshagen T, Espeseth T, Fatouros-Bergman H, Flyckt L, Franke B, Frei O, Haatveit B, Håberg AK, Harbo HF, Hartman CA, Heslenfeld D, Hoekstra PJ, Høgestøl EA, Jernigan TL, Jonassen R, Jönsson EG, Kirsch P, Kłoszewska I, Kolskår KK, Landrø NI, Hellard SL, Lesch K-P, Lovestone S, Lundervold A, Lundervold AJ, Maglanoc LA, Malt UF, Mecocci P, Melle I, Meyer-Lindenberg A, Moberget T, Norbom LB, Nordvik JE, Nyberg L, Oosterlaan J, Papalino M, Papassotiropoulos A, Pauli P, Pergola G, Persson K, Richard G, Rokicki J, Sanders A-M, Selbæk G, Shadrin AA, Smeland OB, Soininen H, Sowa P, Steen VM, Tsolaki M, Ulrichsen KM, Vellas B, Wang L, Westman E, Ziegler GC, Zink M, Andreassen OA, Westlye LT. 2019. Common brain disorders are associated with heritable patterns of apparent aging of the brain. Nat Neurosci. 22:1617–1623.

Kim DR, Bale TL, Epperson CN. 2015. Prenatal Programming of Mental Illness: Current Understanding of Relationship and Mechanisms. Curr Psychiatry Rep. 17:5.

Kirkpatrick B, Messias E, Harvey PD, Fernandez-Egea E, Bowie CR. 2008. Is schizophrenia a syndrome of accelerated aging? Schizophr Bull. 34:1024–1032.

Koch CM, Wagner W. 2011. Epigenetic-aging-signature to determine age in different tissues. Aging (Albany NY). 3:1018–1027.

Kolenic M, Franke K, Hlinka J, Matejka M, Capkova J, Pausova Z, Uher R, Alda M, Spaniel F, Hajek T. 2018. Obesity, dyslipidemia and brain age in first-episode psychosis. Journal of Psychiatric Research. 99:151–158.

Koutsouleris N, Davatzikos C, Borgwardt S, Gaser C, Bottlender R, Frodl T, Falkai P, Riecher-Rössler A, Möller HJ, Reiser M, Pantelis C, Meisenzahl E. 2014. Accelerated brain aging in schizophrenia and beyond: A neuroanatomical marker of psychiatric disorders. Schizophr Bull. 40:1140–1153.

Kowalec K, Hannon E, Mansell G, Burrage J, Ori APS, Ophoff RA, Mill J, Sullivan PF. 2019. Methylation age acceleration does not predict mortality in schizophrenia. Translational Psychiatry. 9:157.

Laursen TM, Nordentoft M, Mortensen PB. 2014. Excess early mortality in schizophrenia. Annu Rev Clin Psychol. 10:425–448.

Laursen TM, Trabjerg BB, Mors O, Børglum AD, Hougaard DM, Mattheisen M, Meier SM, Byrne EM, Mortensen PB, Munk-Olsen T, Agerbo E. 2017. Association of the polygenic risk score for schizophrenia with mortality and suicidal behavior - A Danish population-based study. Schizophrenia Research. 184:122–127.

Le TT, Kuplicki RT, McKinney BA., Yeh HW, Thompson WK, Paulus MP. 2018. A Nonlinear Simulation Framework Supports Adjusting for Age When Analyzing BrainAGE. Frontiers in Aging Neuroscience, 10(October), 1-11.

Levine ME, Lu AT, Quach A, Chen BH, Assimes TL, Bandinelli S, Hou L, Baccarelli AA, Stewart JD, Li Y, Whitsel EA, Wilson JG, Reiner1 AP, Aviv1 A, Lohman K, Liu Y, Ferrucci L, Horvath S. 2018. An epigenetic biomarker of aging for lifespan and healthspan. Aging (Albany NY). 10:573–591.

Li S, Wong EM, Joo JHE, Jung CH, Chung J, Apicella C, Stone J, Dite GS, Giles GG, Southey MC, Hopper JL. 2015. Genetic and Environmental Causes of Variation in the Difference Between Biological Age Based on DNA Methylation and Chronological Age for Middle-Aged Women. Twin Res Hum Genet. 18:720–726.

Li Z, He Y, Ma X, Chen X. 2018. Epigenetic age analysis of brain in major depressive disorder. Psychiatry Res. 269:621–624.

Liang SG, Greenwood TA. 2015. The impact of clinical heterogeneity in schizophrenia on genomic analyses. Schizophrenia Research. 161:490–495.

Lin D, Chen J, Ehrlich S, Bustillo JR, Perrone-Bizzozero N, Walton E, Clark VP, Wang Y-P, Sui J, Du Y, Ho BC, Schulz CS, Calhoun VD, Liu J. 2018. Cross-Tissue Exploration of Genetic and Epigenetic Effects on Brain Gray Matter in Schizophrenia. Schizophr Bull. 44:443–452.

Lowe D, Horvath S, Raj K. 2016. Epigenetic clock analyses of cellular senescence and ageing. Oncotarget. 7:8524–8531.

Lu AT, Xue L, Salfati EL, Chen BH, Ferrucci L, Levy D, Joehanes R, Murabito JM, Kiel DP, Tsai P-C, Yet I, Bell JT, Mangino M, Tanaka T, McRae AF, Marioni RE, Visscher PM, Wray NR, Deary IJ, Levine ME, Quach A, Assimes T, Tsao PS, Absher D, Stewart JD, Li Y, Reiner AP, Hou L, Baccarelli AA, Whitsel EA, Aviv A, Cardona A, Day FR, Wareham NJ, Perry JRB, Ong KK, Raj K, Lunetta KL, Horvath S. 2018. GWAS of epigenetic aging rates in blood reveals a critical role for TERT. Nature Communications. 9:387.

Marioni RE, Shah S, McRae AF, Chen BH, Colicino E, Harris SE, Gibson J, Henders AK, Redmond P, Cox SR, Pattie A, Corley J, Murphy L, Martin NG, Montgomery GW, Feinberg AP, Fallin MD, Multhaup ML, Jaffe AE, Joehanes R, Schwartz J, Just AC, Lunetta KL, Murabito JM, Starr JM, Horvath S, Baccarelli AA, Levy D, Visscher PM, Wray NR, Deary IJ. 2015. DNA methylation age of blood predicts all-cause mortality in later life. Genome Biol. 16:25–12.

Marioni RE, Suderman M, Chen BH, Horvath S, Bandinelli S, Morris T, Beck S, Ferrucci L, Pedersen NL, Relton CL, Deary IJ, Hägg S. 2019. Tracking the epigenetic clock across the human life course: A meta-analysis of longitudinal cohort data. J Gerontol A Biol Sci Med Sci. 74:57–61.

McDade TW, Ryan CP, Jones MJ, Hoke MK, Borja J, Miller GE, Kuzawa CW, Kobor MS. 2019. Genome-wide analysis of DNA methylation in relation to socioeconomic status during development and early adulthood. Am J Phys Anthropol. 169:3–11.

McKinney BC, Lin H, Ding Y, Lewis DA, Sweet RA. 2017. DNA methylation evidence against the accelerated aging hypothesis of schizophrenia, npj Schizophrenia. 3:13.

McKinney BC, Lin H, Ding Y, Lewis DA, Sweet RA. 2018. DNA methylation age is not accelerated in brain or blood of subjects with schizophrenia. Schizophrenia Research. 196:39–44.

Mill J, Tang T, Kaminsky Z, Khare T, Yazdanpanah S, Bouchard L, Jia P, Assadzadeh A, Flanagan J, Schumacher A, Wang S-C, Petronis A. 2008. Epigenomic Profiling Reveals DNA-Methylation Changes Associated with Major Psychosis. Am J Hum Genet. 82:696–711.

Mwangi B, Hasan KM, Soares JC: Prediction of individual subject’s age across the human lifespan using diffusion tensor imaging: a machine learning approach. Neuroimage 2013; 75:58–67.

Nelson PG, Promislow DEL, Masel J. 2019. Biomarkers for aging identified in cross-sectional studies tend to be non-causative. J Gerontol A Biol Sci Med Sci.

Nenadić I, Dietzek M, Langbein K, Sauer H, Gaser C. 2017. BrainAGE score indicates accelerated brain aging in schizophrenia, but not bipolar disorder. Psychiatry Research: Neuroimaging. 266:86–89.

Nguyen TT, Eyler LT, Jeste DV. 2018. Systemic biomarkers of accelerated aging in schizophrenia: A critical review and future directions. Schizophr Bull. 44:398–408.

Okazaki S, Otsuka I, Numata S, Horai T, Mouri K, Boku S, Ohmori T, Sora I, Hishimoto A. 2019. Epigenetic clock analysis of blood samples from Japanese schizophrenia patients, npj Schizophrenia. 5:4.

Olabi B, Ellison-Wright I, McIntosh AM, Wood SJ, Bullmore E, Lawrie SM. 2011. Are there progressive brain changes in schizophrenia? a meta-analysis of structural magnetic resonance imaging studies. Biological Psychiatry. 70:88–96.

Ori APS, Olde Loohuis LM, Guintivano J, Hannon E, Dempster E, St Clair D, Bass NJ, McQuillin A, Mill J, Sullivan PF, Kahn RS, Horvath S, Ophoff RA. 2019. Schizophrenia is characterized by age- and sex-specific effects on epigenetic aging. bioRxiv. 388:86.

Ovenden ES, McGregor NW, Emsley RA, Warnich L. 2018. DNA methylation and antipsychotic treatment mechanisms in schizophrenia: Progress and future directions. Prog Neuropsychopharmacol Biol Psychiatry. 81:38–49.

Palaniyappan L, Das T, Dempster K. 2017. The neurobiology of transition to psychosis: Clearing the cache. JPN 42:294–299.

Papanastasiou E, Gaughran F, Smith S. 2011. Schizophrenia as segmental progeria. J R Soc Med. 104:475–484.

Pashayan N, Reisel D, Widschwendter M. 2016. Integration of genetic and epigenetic markers for risk stratification: Opportunities and challenges. Per Med. 13:93–95.

Paus T, Keshavan M, Giedd JN. 2008. Why do many psychiatric disorders emerge during adolescence? Nat Rev Neurosci. 9:947–957.

Pulver AE, Mulle J, Nestadt G, Swartz KL, Blouin JL, Dombroski B, Liang KY, Housman DE, Kazazian HH, Antonarakis SE, Lasseter VK, Wolyniec PS, Thornquist MH, McGrath JA. 2000. Genetic heterogeneity in schizophrenia: Stratification of genome scan data using co-segregating related phenotypes. Mol Psychiatry. 5:650–653.

Ranlund S, Rosa MJ, de Jong S, Cole JH, Kyriakopoulos M, Fu CHY, Mehta MA, Dima D. 2018. Associations between polygenic risk scores for four psychiatric illnesses and brain structure using multivariate pattern recognition. Neuroimage Clin. 20:1026–1036.

Ripke S, Schizophrenia Working Group of the Psychiatric Genomics Consortium. 2014. Biological insights from 108 schizophrenia-associated genetic loci. Nature. 511:421–427.

Roiz-Santiañez R, Suarez-Pinilla P, Crespo-Facorro B. 2015. Brain Structural Effects of Antipsychotic Treatment in Schizophrenia: A Systematic Review. CN. 13:422–434.

Sabunciyan S. 2019. Gene Expression Profiles Associated with Brain Aging are Altered in Schizophrenia. Sci Rep. 9:5896.

Schnack HG, van Haren NEM, Nieuwenhuis M, Hulshoff Pol HE, Cahn W, Kahn RS. 2016. Accelerated brain aging in schizophrenia: A longitudinal pattern recognition study. AJP. 173:607–616.

Shahab S, Mulsant BH, Levesque ML, Calarco N, Nazeri A, Wheeler AL, Foussias G, Rajji TK, Voineskos AN. 2019. Brain structure, cognition, and brain age in schizophrenia, bipolar disorder, and healthy controls. Neuropsychopharmacology. 44:898–906.

Shivakumar V, Kalmady SV, Venkatasubramanian G, Ravi V, Gangadhar BN. 2014. Do schizophrenia patients age early? Asian J Psychiatr. 10:3–9.

Soreq L, Rose J, Soreq E, Hardy J, Trabzuni D, Cookson MR, Smith C, Ryten M, Patani R, Ule J. 2017. Major Shifts in Glial Regional Identity Are a Transcriptional Hallmark of Human Brain Aging. Cell Rep. 18:557–570.

Swathy B, Banerjee M. 2017. Understanding epigenetics of schizophrenia in the backdrop of its antipsychotic drug therapy. Epigenomics. 9:721–736.

Tang B, Chang W-L, Lanigan CM, Dean B, Sutcliffe JG, Thomas EA. 2009. Normal human aging and early-stage schizophrenia share common molecular profiles. Aging Cell. 8:339–342.

Valiathan R, Ashman M, Asthana D. 2016. Effects of Ageing on the Immune System: Infants to Elderly. Scand J Immunol. 83:255–266.

Valizadeh SA, Hanggi J, Merillat S, Jancke L. 2017. Age prediction on the basis of brain anatomical measures. Hum Brain Mapp. 38:997–1008.

van Dongen J, Nivard MG, Willemsen G, Hottenga J-J, Helmer Q, Dolan CV, Ehli EA, Davies GE, van Iterson M, Breeze CE, Beck S, Suchiman HE, Jansen R, van Meurs JB, Heijmans BT, Slagboom PE, Boomsma DI. 2016. Genetic and environmental influences interact with age and sex in shaping the human methylome. Nature Communications. 7:11115.

Van Gestel H, Franke K, Petite J, Slaney C, Garnham J, Helmick C, Johnson K, Uher R, Alda M, Hajek T. Brain age in bipolar disorders: Effects of lithium treatment. Australian & New Zealand Journal of Psychiatry. 2019 Dec;53(12):1179-88.

van Haren NEM, Hulshoff Pol HE, Schnack HG, Cahn W, Mandl RCW, Collins DL, Evans AC, Kahn RS. 2007. Focal gray matter changes in schizophrenia across the course of the illness: a 5-year follow-up study. Neuropsychopharmacology 32:2057–2066.

van Haren NEM, Hulshoff Pol HE, Schnack HG, Cahn W, Brans R, Carati I, Rais M, Kahn RS. 2008. Progressive Brain Volume Loss in Schizophrenia Over the Course of the Illness: Evidence of Maturational Abnormalities in Early Adulthood. Biological Psychiatry. 63:106–113.

Viana J, Hannon E, Dempster E, Pidsley R, Macdonald R, Knox O, Spiers H, Troakes C, Al-Saraj S, Turecki G, Schalkwyk LC, Mill J. 2017. Schizophrenia-associated methylomic variation: molecular signatures of disease and polygenic risk burden across multiple brain regions. Hum Mol Genet. 26:210–225.

Voisey J, Lawford BR, Morris CP, Wockner LF, Noble EP, Young RMD, Mehta D. 2017. Epigenetic analysis confirms no accelerated brain aging in schizophrenia, npj Schizophrenia. 3:26.

Walton E, Hass J, Liu J, Roffman JL, Bernardoni F, Roessner V, Kirsch M, Schackert G, Calhoun V, Ehrlich S. 2016. Correspondence of DNA methylation between blood and brain tissue and its application to schizophrenia research. Schizophr Bull. 42:406–414.

Weidner CI, Lin Q, Koch CM, Eisele L, Beier F, Ziegler P, Bauerschlag DO, Jöckel K-H, Erbel R, Mühleisen TW, Zenke M, Brümmendorf TH, Wagner W. 2014. Aging of blood can be tracked by DNA methylation changes at just three CpG sites. Genome Biol. 15:R24.

Wolf EJ, Maniates H, Nugent N, Maihofer AX, Armstrong D, Ratanatharathorn A, Ashley-Koch AE, Garrett M, Kimbrel NA, Lori A, VA Mid-Atlantic MIRECC Workgroup, Aiello AE, Baker DG, Beckham JC, Boks MP, Galea S, Geuze E, Hauser MA, Kessler RC, Koenen KC, Miller MW, Ressler KJ, Risbrough V, Rutten BPF, Stein MB, Ursano RJ, Vermetten E, Vinkers CH, Uddin M, Smith AK, Nievergelt CM, Logue MW. 2018. Traumatic stress and accelerated DNA methylation age: A meta-analysis. Psychoneuroendocrinology. 92:123–134.

Zhang Q, Vallerga CL, Walker RM, Lin T, Henders AK, Montgomery GW, He J, Fan D, Fowdar J, Kennedy M, Pitcher T, Pearson J, Halliday G, Kwok JB, Hickie I, Lewis S, Anderson T, Silburn PA, Mellick GD, Harris SE, Redmond P, Murray AD, Porteous DJ, Haley CS, Evans KL, McIntosh AM, Yang J, Gratten J, Marioni RE, Wray NR, Deary IJ, McRae AF, Visscher PM. 2018. Improved prediction of chronological age from DNA methylation limits it as a biomarker of ageing. bioRxiv. 80:245.

